# Identification of spatial variations in COVID-19 epidemiological data using K-Means clustering algorithm: a global perspective

**DOI:** 10.1101/2020.06.03.20121194

**Authors:** Viswa Chaitanya Chandu

## Abstract

**Background:** Discerning spatial variations of COVID-19 through quantitative analysis operating on the geographically designated datasets relating to socio-demographics and epidemiological data facilitate strategy planning in curtailing the transmission of the disease and focus on articulation of necessary interventions in an informed manner.

**Methods:** K-means clustering was employed on the available country-specific COVID-19 epidemiological data and the influential background characteristics. Country-specific case fatality rates and the average number of people tested positive for COVID-19 per every 10,000 population in each country were derived from the WHO COVID-19 situation report 107, and were used for clustering along with the background characteristics of proportion of country’s population aged >65 years and percentage GDP spent as public health expenditure.

**Results:** The algorithm grouped the 89 countries into cluster ‘1’ and Cluster ‘2’ of sizes 54 and 35, respectively. It is apparent that Americas, European countries, and Australia formed a major part of cluster ‘2’ with high COVID-19 case fatality rate, higher proportion of country’s population tested COVID-19 positive, higher percentage of GDP spent as public health expenditure, and greater percentage of population being more than 65 years of age.

**Conclusion:** In spite of the positive correlation between high public health expenditure (%GDP) and COVID-19 incidence, case fatality rate, the immediate task ahead of most of the low and middle income countries is to strengthen their public health systems realizing that the correlation found in this study could be spurious in light of the underreported number of cases and poor death registration.

## Introduction

Since its early origins in late December, 2019, Coroanvirus disease (COVID-19) has steadily pushed the world into huge health and economic crisis. On 30^th^ January, 2020, COVID-19 has been identified by World Health Organization as a public health emergency of international concern, and was declared as a pandemic on 11^th^ March, 2020.^1^ According to the World Health Organization’s situation report 107 on 6^th^ May, 2020, an unprecedented number of 181 countries across the globe reported laboratory confirmed COVID-19 positive cases.^2^ A close observation of the global data on COVID-19 positive cases and deaths associated with COVID-19 reveals an apparent distinction between nations. Discerning spatial variations of COVID-19 through quantitative analysis operating on the geographically designated datasets relating to socio-demographics and epidemiological data facilitate strategy planning in curtailing the transmission of the disease and focus on articulation of necessary interventions in an informed manner.

## Methods

In order to understand the spatial variations, we have employed K-means clustering on the available country-specific COVID-19 epidemiological data and the influential background characteristics. In this study, only those countries with at least 1000 confirmed positive cases were included, which is 89 according to the WHO COVID-19 situation report 107. K-means clustering is a robust, extensively used clustering algorithm which results in the identification of K non-overlapping clusters.^3^ Two COVID-19 epidemiological variables and two influential background characteristics were selected as attributes in this study. The COVID-19 epidemiological data derived from the WHO COVID-19 situation report 107 include country-specific case fatality rates and the average number of people tested positive for COVID-19 per every 10,000 population in each country. The influential background characteristics chosen were the percentage of country’s population aged 65 and more and the percentage of GDP spent towards public health expenditure in the country.^4^ After selecting the aforementioned attributes to be used in the partitioning algorithm, K-means clustering analysis was performed using SPSS version 20 software (IBM SPSS statistics for windows version 20, Armonk, NY, USA). Since the four attributes used are measured on different scales, standardized scores were obtained for all the attributes and used for K-means clustering. As the desired number of clusters in K-means clustering is user specified, elbow method was employed in determination of optimum number of clusters to ensure objectivity. The algorithm was quantified with different values of K (1<K≤5), and the line diagram plotted for within cluster sum of squares demonstrated a sharp bend at K=2. This observation of the optimum number of clusters to be 2 was supported by the dendrogram obtained from hierarchical clustering algorithm as well. K-means clustering with K=2 was performed, and it took 4 iterations for the data points to stably cluster into 2 groups with the initial randomly selected centroids moved to the true centroids of the clusters.

## Results

The algorithm grouped the 89 countries into cluster ‘1’ and Cluster ‘2’ of sizes 54 and 35, respectively. The Euclidean distance between final cluster centers was 2.634. Figure 1 shows the geographical clustering of countries on the world map based on the attributes of interest plotted using the QGIS 3.1.2.2 software.^5^ It is apparent that Americas, European countries, and Australia formed a major part of cluster ‘2’ with high COVID-19 case fatality rate, higher proportion of country’s population tested COVID-19 positive, higher percentage of GDP spent as public health expenditure, and greater percentage of population being more than 65 years of age. Figure 2 shows the matrix scatter plot of the correlation between standardized values of the study attributes stratified by cluster. The clear distinction between the two cluster centroids in the correlation matrix demonstrate the within cluster homogeneity and inter-cluster heterogeneity. Independent samples t-test was employed to check the inter-cluster difference in the distance of cluster members from their corresponding cluster centers (t =-3.96; p = 0.001).

**Figure 1:**
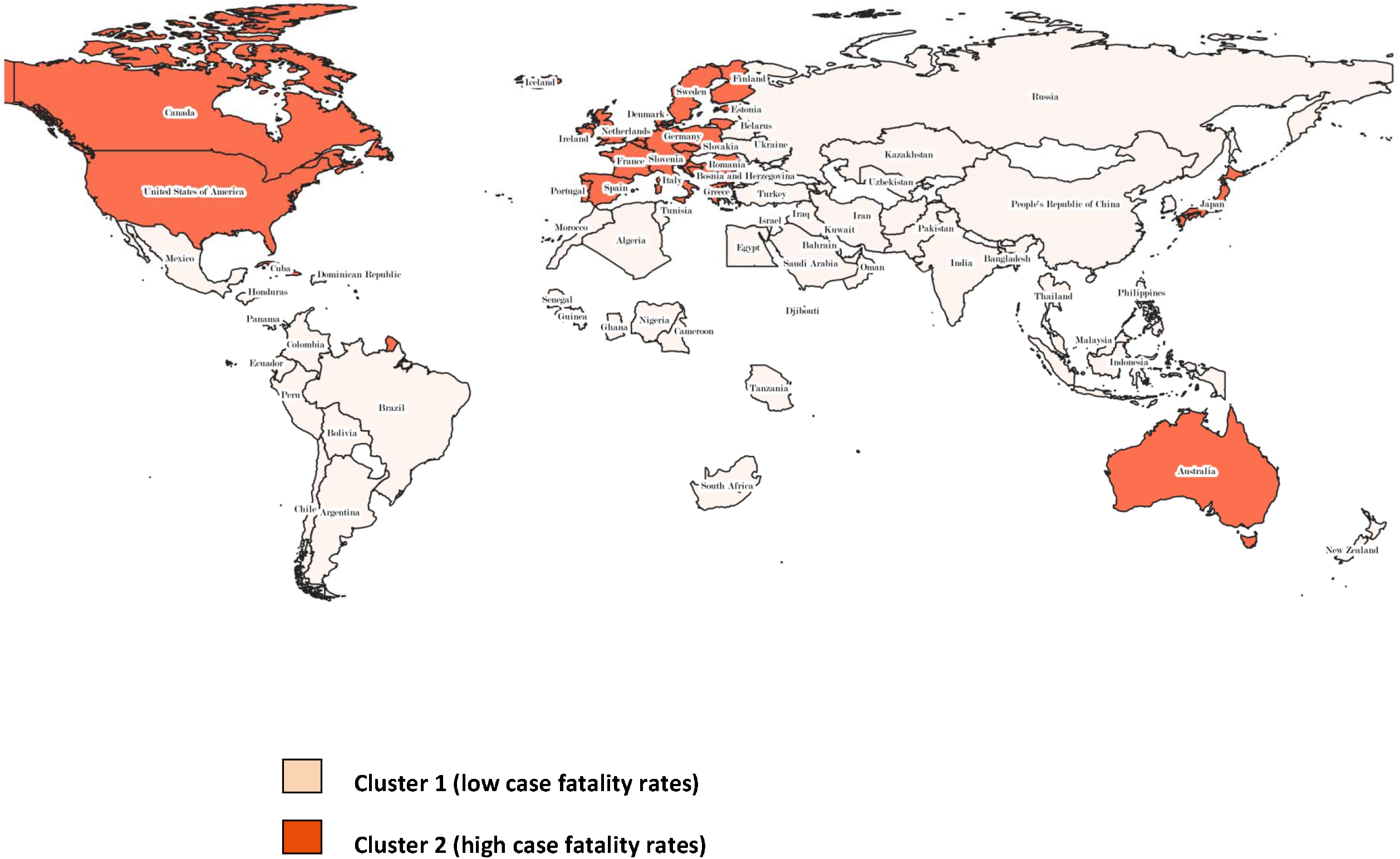
Pictorial representation of the geographical clustering based on the study attributes.

**Figure 2:**
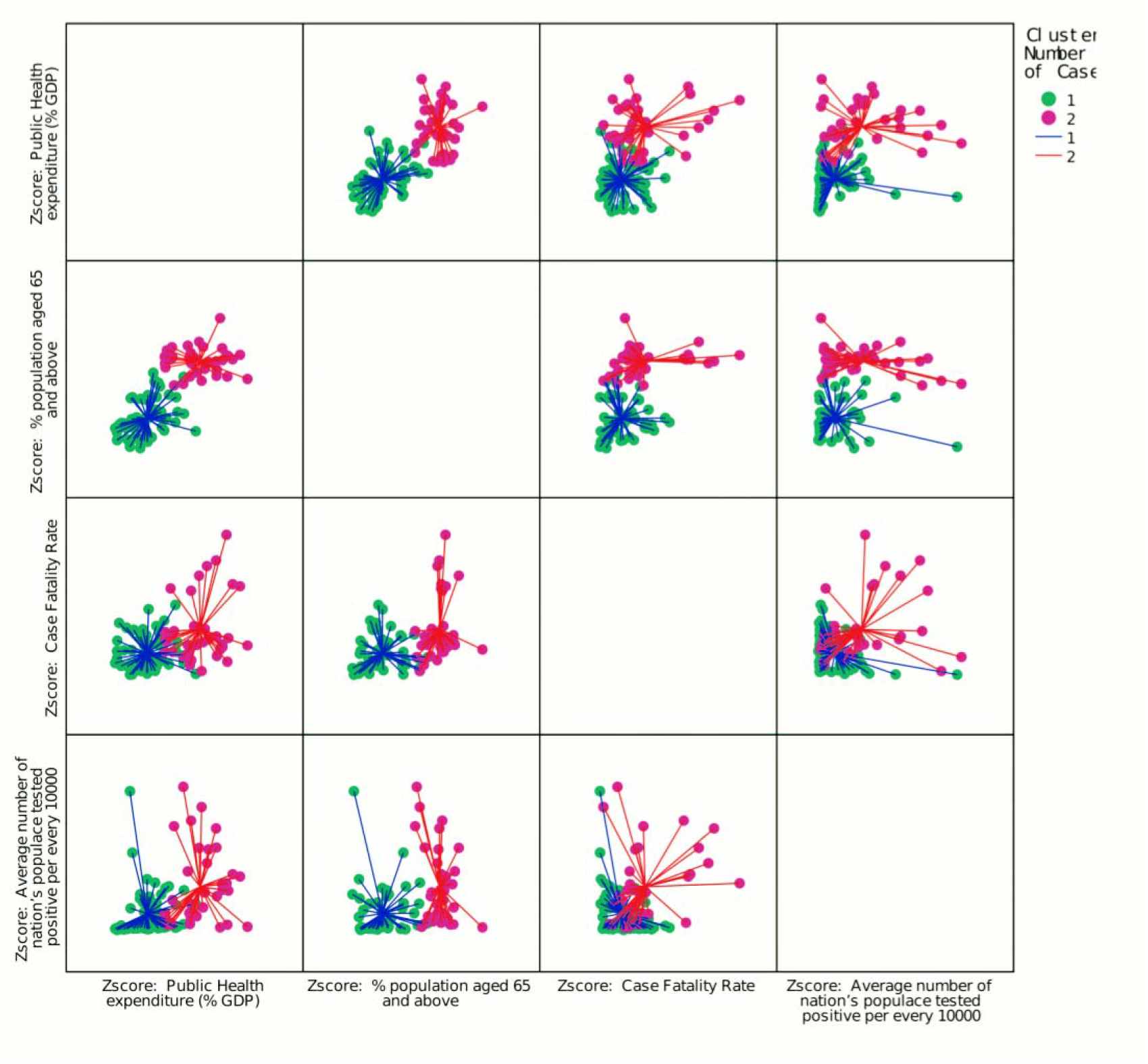
Matrix scatter plot of the correlation between standardized values of the study attributes stratified by cluster.

## Discussion

The study results highlight a rather unanticipated observation that the COVID-19 case fatality rate and the average number per 10,000 population tested COVID-19 positive were more in those countries with high public health expenditure (% GDP) which could be considered a proxy measure for stronger public health systems in place. However, the rationale for the observation of low case fatality rate among countries in cluster ‘1’ could be incomplete death registration rates with relatively weaker public health systems in place. Even there is limited availability of COVID-19 test kits in these countries decreasing the number of people tested per day, which consequently reduces the proportion of country’s population positively tested for COVID-19. Therefore, in spite of the positive correlation between high public health expenditure (%GDP) and COVID-19 incidence, case fatality rate, the immediate task ahead of most of the low and middle income countries is to strengthen their public health systems realizing that the correlation found in this study could be spurious in light of the underreported number of cases and poor death registration.^6^ The underlying co morbidities for COVID-19 positive persons, though would influence case fatality rates, could not be integrated in the present analysis because of the non availability of person level data at this stage. Nevertheless, the study surfaces an important COVID-19 spatial pattern recognition and provides an argument for the statistics of some countries to be under reported. The negative consequences of under reporting could be false assurance among the public and also carry the risk of reinforcing myths among the public that people from certain geographic regions are innately more resistant.

## Data Availability

Data is available

## References

1) World Health Organization. WHO Director-General’s opening remarks at the media briefing on COVID-19 - 11 March 2020. https://www.who.int/dg/speeches/detail/who-director-general-sopening-remarks-at-the-media-briefing-on-covid-19-11-march-2020.

2) World Health Organization. 2020. b. Coronavirus disease 2019 (COVID-19): situation report-107 [accessed on 6th May, 2020]. Retrieved from: https://www.who.int/docs/default-source/coronaviruse/situation-reports/20200506covid-19-sitrep-107.pdf?sfvrsn=159c3dc_2

3) Junjie Wu. 2012. Advances in K-means Clustering: A Data Mining Thinking. Springer Publishing Company, Incorporated.

4) Xu K, Soucat A & Kutzin J et al. Public Spending on Health: A Closer Look at Global Trends. Geneva: World Health Organization; 2018 (WHO/HIS/HGF/HFWorkingPaper/18.3). Licence: CC BY-NC-SA 3.0 IGO.

5) QGIS Development Team (2020). QGIS Geographic Information System. Open Source Geospatial Foundation Project. http://qgis.osgeo.org

6) Glassman A, Chalkidou K, Sullivan R. Does one size fits all? Realistic alternatives for COVID-19 response in low income countries. Center for Global development. 2022 April 2. Retrieved from: https://www.cgdev.org/blog/does-one-size-fit-all-realistic-alternatives-covid-19-response-low-income-countries

